# The Flexor Carpi Ulnaris Cross-Sectional Area is a Potential Early Indicator of ICU-Acquired Weakness: A Prospective Cohort Study

**DOI:** 10.64898/2026.02.09.26345643

**Authors:** Keiichiro Aoki, Fumihito Kasai, Kazuki Komaba, Jin Saito, Akira Yoshikawa, Naonori Tashiro, Hiroyasu Inoue, Kengo Uchibori, Miwa Fukazawa

## Abstract

**Background:** In critically ill patients admitted to the intensive care unit (ICU), rapid skeletal muscle atrophy frequently develops in the acute phase. This ICU-acquired weakness can significantly impair long-term physical function. Although the biceps brachii cross-sectional area (CSA) is commonly used to assess muscle atrophy, its ultrasound imaging can be technically challenging, and the flexor carpi ulnaris may offer a more accessible alternative. Therefore, this study aimed to investigate whether CSA changes of the flexor carpi ulnaris correlate with those of the biceps brachii in critically ill patients admitted to the ICU, as well as whether the flexor carpi ulnaris CSA reflects systemic muscle atrophy in the acute phase of the ICU stay.

**Methods:** Twenty critically ill patients admitted to the ICU underwent serial ultrasound assessment of the biceps brachii and flexor carpi ulnaris CSAs on days 0, 5, 7, and 14 after admission. Longitudinal changes in CSA were analyzed using the Friedman and Wilcoxon signed-rank tests. Correlations between the biceps brachii and flexor carpi ulnaris were examined using Spearman’s rank correlation, and structural equation modeling was applied to explore causal relationships between clinical variables and CSA changes.

**Results:** Significant CSA reductions were observed in both the flexor carpi ulnaris (−20.6%) and biceps brachii (−16.3%) by day 14, and the relative CSA changes of the biceps brachii and flexor carpi ulnaris showed a moderate positive correlation (ρ = 0.5489, p = 0.0122). Structural equation modeling analysis revealed that the biceps brachii CSA change had positive effect on that of the flexor carpi ulnaris (β = 0.249, p = 0.0011). Moreover, body mass index was positively associated with the baseline flexor carpi ulnaris CSA (β = 0.042, p = 0.0004). However, the baseline flexor carpi ulnaris CSA was not a significant predictor of subsequent CSA changes.

**Conclusion:** Ultrasound measurement of the flexor carpi ulnaris CSA offers a practical alternative to that of the biceps brachii for early detection of muscle wasting in ICU patients. Given its anatomical accessibility and high sensitivity to early atrophic changes, it may serve as a feasible screening tool for ICU-acquired weakness and inform timely interventions.

## Background

In critically ill patients admitted to the intensive care unit (ICU), rapid progression of skeletal muscle loss and muscle weakness is known to occur from the acute phase—these conditions are recognized as key components of post-intensive care syndrome [1]. In particular, ICU-acquired weakness (ICU-AW)—defined as muscle weakness newly developed during the ICU stay—has been reported to have long-term adverse effects on physical function and health-related quality of life after discharge from the ICU [2, 3].

ICU-AW is commonly assessed using the Medical Research Council (MRC) score, administered by healthcare professionals such as physical and occupational therapists involved in early rehabilitation [4]. However, the assessment requires patient wakefulness and cooperation, which often makes it difficult to perform in the early ICU phase [5]. Therefore, ultrasound-based measurement of muscle cross-sectional area (CSA) has attracted attention as a more objective and non-invasive method for assessing muscle mass [6].

Traditionally, muscles such as the rectus femoris and biceps brachii (BB) have been used for muscle mass evaluation. For example, Puthucheary et al. [7] reported that in ICU patients on mechanical ventilation, the CSA of the rectus femoris decreased by an average of 17.7% after 10 days of ICU admission. Similarly, in another study, the BB was shown to atrophy by approximately 13–17% within 7 days [8]. These muscles are regarded as representative sites that reflect overall muscle wasting; however, in clinical practice, some challenges to muscle mass evaluation using these muscles exist. For example, the rectus femoris is located relatively deep, and visualization of the BB can be difficult depending on patient positioning and ultrasound conditions, requiring skilled techniques for reliable measurements [9]. Furthermore, access to the lower limbs or upper arms may be restricted in patients undergoing intensive interventions such as extracorporeal membrane oxygenation, intra-aortic balloon pumping, or continuous hemodiafiltration, all of which involve vascular access and external devices [10]. These limitations indicate that conventional muscle sites may not always be easily assessable at the bedside.

Given these issues, we focused on the flexor carpi ulnaris (FCU), a superficial forearm muscle, as a potential new, simple, and reproducible target for muscle mass assessment. The FCU is located in the superficial layer and is relatively easy to visualize using ultrasound, even in the flexed elbow position; in fact, it is reportedly more clearly visualized than other forearm muscles [11]. Moreover, the forearm is generally accessible through postural adjustments or upper limb elevation, offering high flexibility for measurement in ICU settings—a significant clinical advantage [12].

Therefore, in this study, we longitudinally measured the CSA of the FCU using ultrasound in critically ill patients admitted to the ICU and examined its relationship with the CSA of the BB, a conventional assessment site. We then explored the potential of the FCU CSA as a useful index for objectively capturing muscle wasting, even in severely ill patients, for whom conventional motor function assessments are difficult.

## Methods

### Study Design and Ethics

This prospective observational study was conducted at two tertiary care hospitals in Japan between 2023 and 2025. The study was conducted in accordance with the principles of the Declaration of Helsinki, and the protocol was approved by the Institutional Review Board of Showa Medical University (Approval No. 2023-041-B).

As patients in the ICU are often in a critical condition upon admission, obtaining written informed consent in advance was frequently impractical. Therefore, considering that the ultrasound measurements were part of routine clinical care, verbal informed consent for study participation was obtained from the patient or their legally authorized representative (e.g., family member) once the patient’s condition stabilized or communication became feasible. The content of the consent was documented in the medical records. Since ultrasound measurements were performed as part of standard care, consent was occasionally obtained 1–2 days after the initial measurements.

### Participants

The inclusion criteria were as follows: (1) patients registered within 24 hours of ICU admission, (2) patients judged by the attending physician to require ≥48 hours of mechanical ventilation, (3) patients with a predicted ICU stay of ≥5 days, and (4) patients aged ≥18 years. Patients were excluded from the study according to the following exclusion criteria: (1) presence of limb trauma or amputation, (2) diagnosis of primary neuromuscular disease, or (3) inability to acquire clear ultrasound images for measurement.

### Sample Size

Sample size estimation was based on statistical power considerations for the Friedman test and Spearman’s rank correlation coefficient. According to the theoretical framework of G*Power developed by Faul et al. [13], a moderate effect size (f = 0.25), power = 0.80, and α = 0.05 would require a minimum of 20 participants for adequate power using the Friedman test. For correlation analysis of CSA, Bonett and Wright [14] recommended a sample size of 18–25 cases to detect a moderate correlation (ρ = 0.5–0.6). Therefore, to improve estimation precision and take potential dropouts into account, a target sample size of 25 participants was set.

### Ultrasound Measurement Protocol

CSA was measured using a SONIMAGE MX1 ultrasound system (Konica Minolta, Tokyo, Japan) with a linear probe (L11-3). Ample contact gel was used to avoid compression of the muscle by the probe, which was consistently placed perpendicular to the longitudinal axis of the limb.

The measurement sites and postures were as follows: the FCU was measured at the midpoint between the olecranon and ulnar styloid, with the patient lying in a supine position, the elbow flexed at 90°, and the forearm in a neutral position; and the BB was measured at the point two-thirds distal along the line from the acromion to the cubital fossa crease, with the patient lying in a supine position with the elbow extended.

After clear ultrasound images were obtained, the CSA was calculated by tracing the outer boundary of the muscle belly (Figure 1a, 1b). To ensure reproducibility, measurement sites were marked on the skin and all measurements were taken from the same site. Each muscle was measured twice consecutively by a single evaluator, and the mean value was used for analysis.

**Figure 1.**
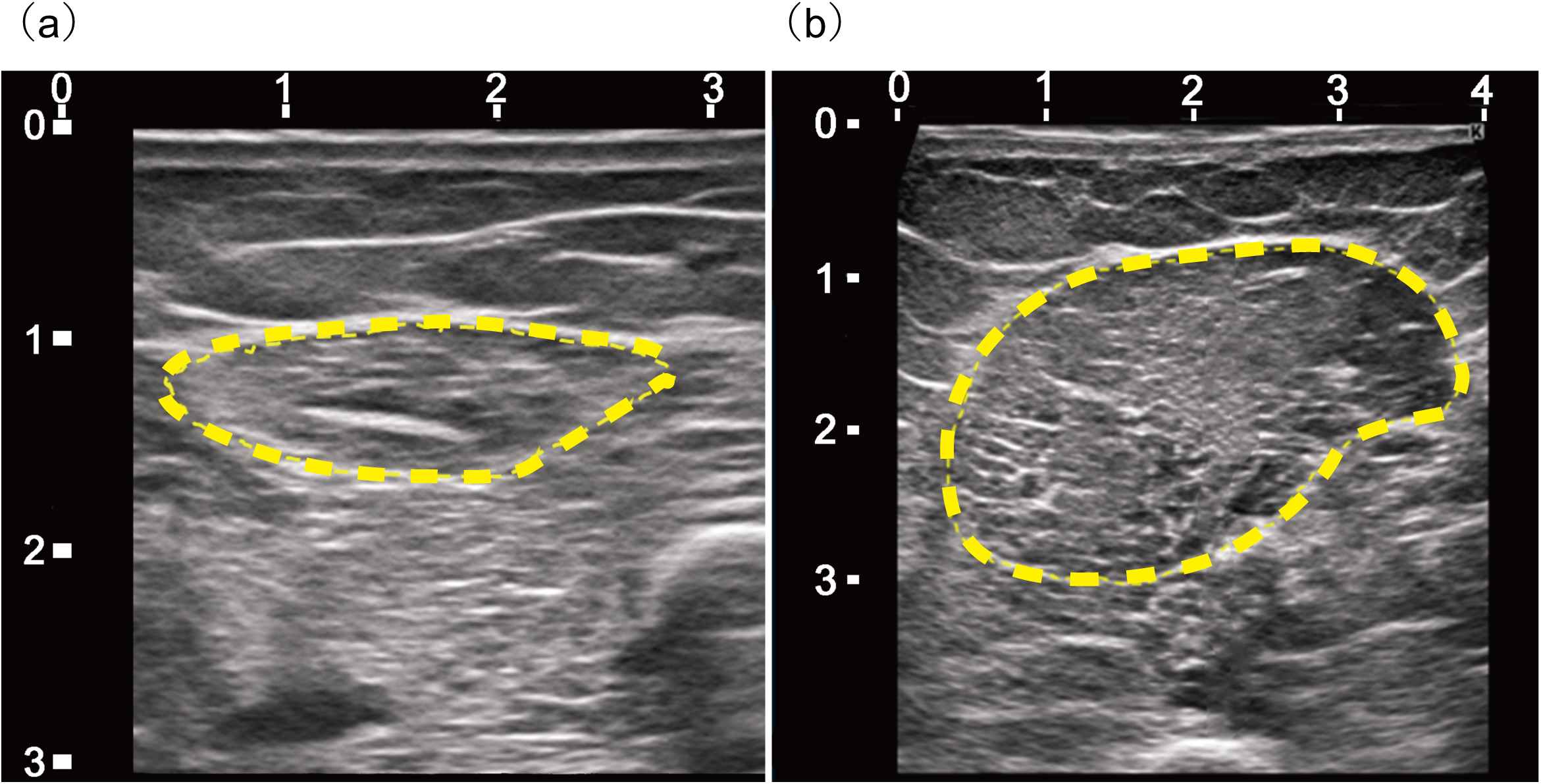
Representative Ultrasound Images of the Flexor Carpi Ulnaris and Biceps Brachii Muscles. (a) Cross-sectional ultrasound image of the flexor carpi ulnaris. (b) Cross-sectional ultrasound image of the biceps brachii. The muscle cross-sectional area was calculated by manually tracing the outer contour of the muscle belly (yellow dashed line) when its boundaries were clearly visible on the image.

### Reliability of Ultrasound Measurements

As this multicenter study was conducted at two hospitals, it was necessary to confirm the reliability and compatibility of the CSA measurements obtained by Examiner A and Examiner B from each facility. Therefore, prior to the main study, the inter-rater reliability of the ultrasound measurements was tested in five healthy adults (three men and two women, aged 24–36 years, mean age 29.2 ± 4.8 years). Both the FCU and BB were measured twice by each of the two examiners (A and B) to evaluate the inter-rater reliability.

### Evaluation Items and Outcomes

The primary outcome of this study was the longitudinal change in the CSA of the FCU and BB during the ICU stay. Measurements were performed at four time points: day 0 (within 24 hours of ICU admission), day 5, day 7, and day 14. These time points were set with reference to the assessment schedule used by Zhang et al. [15], adjusted for feasibility and clinical workflow at the two participating institutions.

The secondary outcomes of this study included the clinical severity and course, rehabilitation-related factors, and discharge outcomes. These three outcomes were exploratorily examined for association with the following indicators: (1) clinical severity and course: ICU length of stay, total hospital days, Sequential Organ Failure Assessment (SOFA) score, duration of mechanical ventilation, and Richmond Agitation-Sedation Scale; (2) rehabilitation-related factors: days to rehabilitation initiation, Functional Status Score for the ICU (FSS-ICU), Barthel Index (BI), and ICU Mobility Scale (IMS); and (3) discharge outcomes: discharge to home, transfer, or death. These secondary data were extracted from medical records. The FSS-ICU, BI, and IMS scores were collected within one week of rehabilitation initiation.

### Statistical Analysis

All statistical analyses were performed using JMP Pro (Version 16, SAS Institute Inc., Cary, NC, USA), with statistical significance set at p < 0.05.

#### Primary Analysis

Longitudinal changes in the CSA of the FCU and BB were assessed at four time points: day 0 (baseline), day 5, day 7, and day 14. As the repeated measurements were not normally distributed, the Friedman test was used. When significant differences were found, pairwise comparisons between time points were conducted using the Wilcoxon signed-rank test. Moreover, relative percentage changes in CSA from day 0 to each subsequent time point were calculated.

#### Correlation Analysis Between the FCU and BB

To assess whether the FCU has the potential to serve as a surrogate marker for BB muscle mass, two correlation analyses were performed. First, Spearman’s rank correlation coefficient was calculated for absolute CSA values of the FCU and BB at day 0 to examine their relationship upon ICU admission. Second, the correlation between relative percentage changes in the FCU and BB CSAs from days 0–14 was calculated to evaluate whether atrophic trends in both muscles were synchronized.

#### Secondary Analysis

To explore factors influencing muscle mass changes after ICU admission and examine how CSA changes in the FCU relate to those in the BB, path analysis using structural equation modeling (SEM) was performed. The design of the structural model and path settings were based on previous reports [7, 9, 16] regarding the early onset of muscle atrophy in the ICU and the impact of clinical severity and immobilization on muscle mass. Following the theoretical SEM framework described by Hooper et al. [17], the model included observed variables such as age, body mass index (BMI), SOFA score, and duration of mechanical ventilation, as well as baseline CSA (FCU0 and BB0) and the CSA percentage changes over 14 days (FCU 0–14-day change%, BB 0–14-day change%). The model included self-predictive paths (e.g., FCU0 → FCU 0–14-day change%), a path linking changes between different muscles (BB 0–14-day change% → FCU 0–14-day change%), and paths from clinical background factors to the FCU 0–14-day change%. Latent variables were not included in this model.

## Results

### Reliability of Ultrasound Measurements

Summary statistics and intraclass correlation coefficients (ICCs) from the inter- and intra-rater reliability testing of CSA measurement using ultrasound are presented in Tables 1 and 2. The inter-rater ICC for measurement of the BB CSA was 0.96, and the intra-rater ICCs for Examiners A and B were 0.97 and 0.98, respectively, indicating excellent reliability. Similarly, for the FCU CSA, the inter-rater ICC was 0.98 and the intra-rater ICCs were 0.99 for both Examiners A and B, demonstrating very high reproducibility. Based on this reliability, ultrasound measurement of the participants’ BB and FCU CSAs were conducted by both Examiners A and B.

**Table 1.** Summary Statistics for Inter-Rater Reliability Testing of CSA Measurement Using Ultrasound.

| Participant | Age<br>(Years) | Sex | Flexor Carpi Ulnaris |  | Biceps Brachii |  |
| --- | --- | --- | --- | --- | --- | --- |
|  |  |  | Mean<br>(cm<sup>2</sup>) | SD | Mean<br>(cm<sup>2</sup>) | SD |
| No. 1 | 26-30 | Male | 1.59 | 0.01 | 8.00 | 0.76 |
| No. 2 | 21-25 | Female | 0.91 | 0.01 | 4.45 | 0.22 |
| No. 3 | 21-25 | Female | 1.14 | 0.03 | 5.09 | 0.66 |
| No. 4 | 36-40 | Male | 1.46 | 0.04 | 7.65 | 0.75 |
| No. 5 | 31-35 | Male | 1.14 | 0.05 | 6.00 | 0.66 |

Abbreviations: SD, standard deviation

**Table 2.**
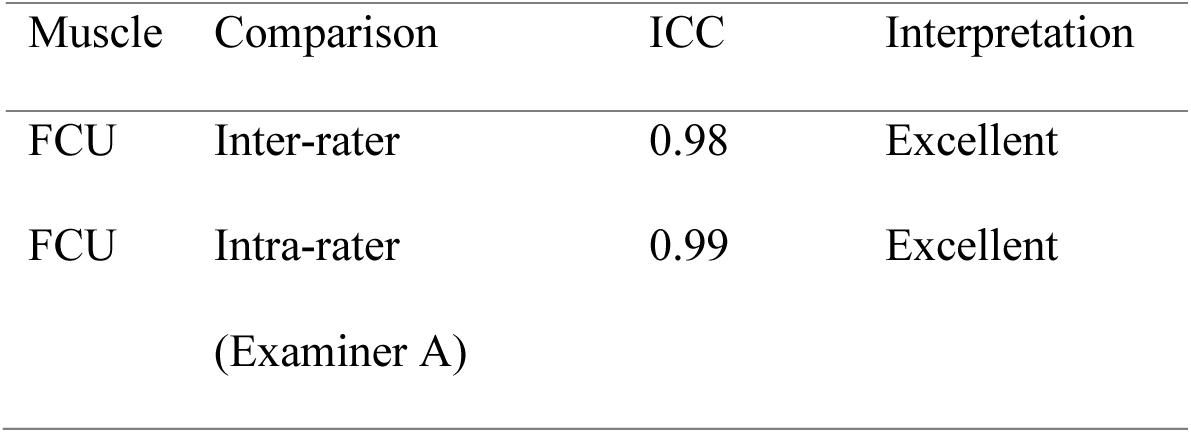
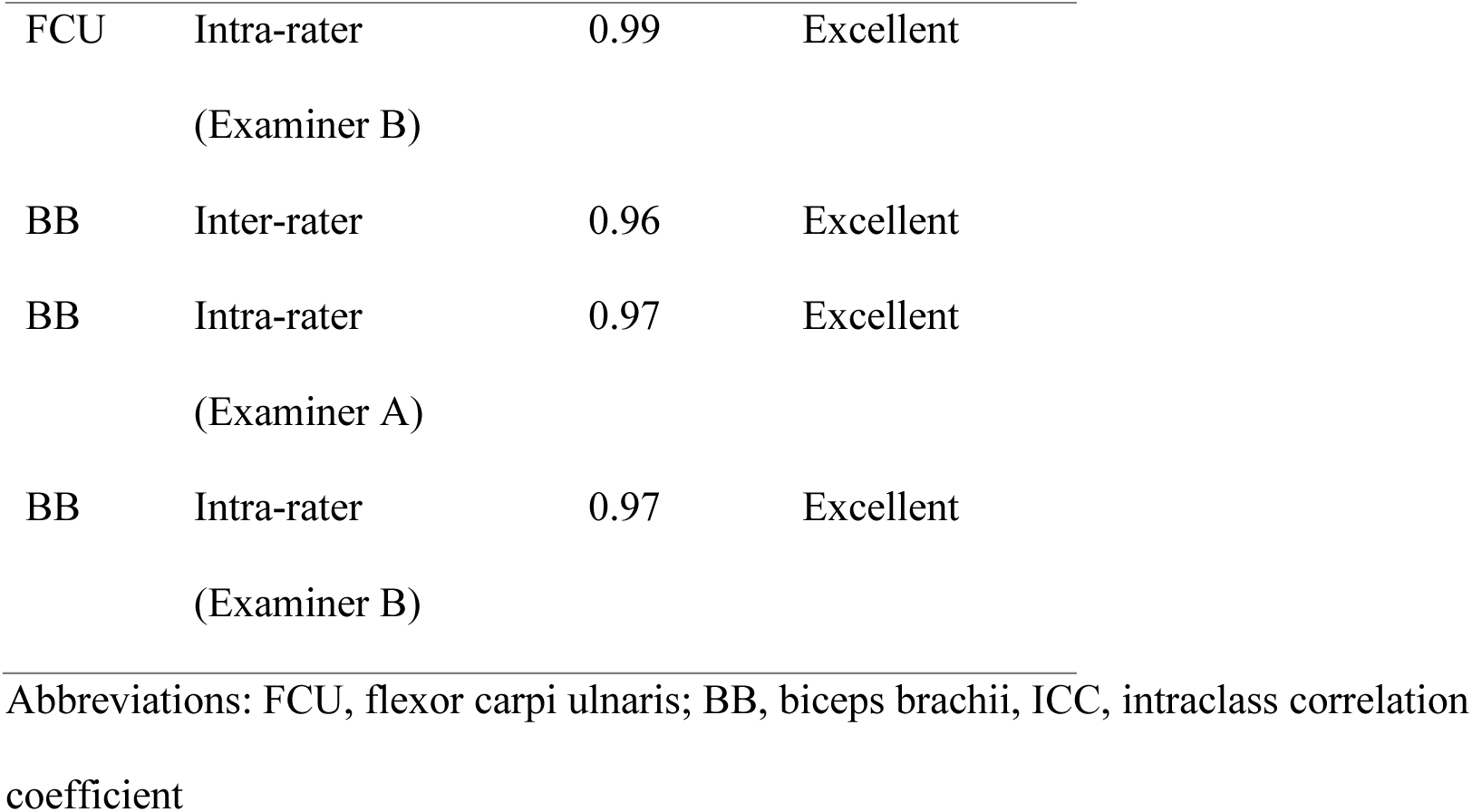
ICCs for the Ultrasound Measurement of the FCU and BB CSAs.

| Muscle | Comparison | ICC | Interpretation |
| --- | --- | --- | --- |
| FCU | Inter-rater | 0.98 | Excellent |
| FCU | Intra-rater<br>(Examiner A) | 0.99 | Excellent |
| FCU | Intra-rater<br>(Examiner B) | 0.99 | Excellent |
| BB | Inter-rater | 0.96 | Excellent |
| BB | Intra-rater<br>(Examiner A) | 0.97 | Excellent |
| BB | Intra-rater<br>(Examiner B) | 0.97 | Excellent |

Abbreviations: FCU, flexor carpi ulnaris; BB, biceps brachii, ICC, intraclass correlation coefficient

### Participant Characteristics

Although the target sample size was 25 participants, only 23 were enrolled within the study period. Of these, three participants were excluded owing to unclear ultrasound images or insufficient data across all time points. Therefore, a total of 20 participants (10 males, 10 females) were included in the final analysis, with a median age of 57 (range 33–82) years. Disease categories included infectious/septic conditions, neurological disorders/post-cardiopulmonary arrest, cardiovascular diseases, and respiratory/metabolic disorders. The median SOFA score was 9 (0–15), and the median duration of mechanical ventilation was 23 (5–70) days (Table 3).

**Table 3.**
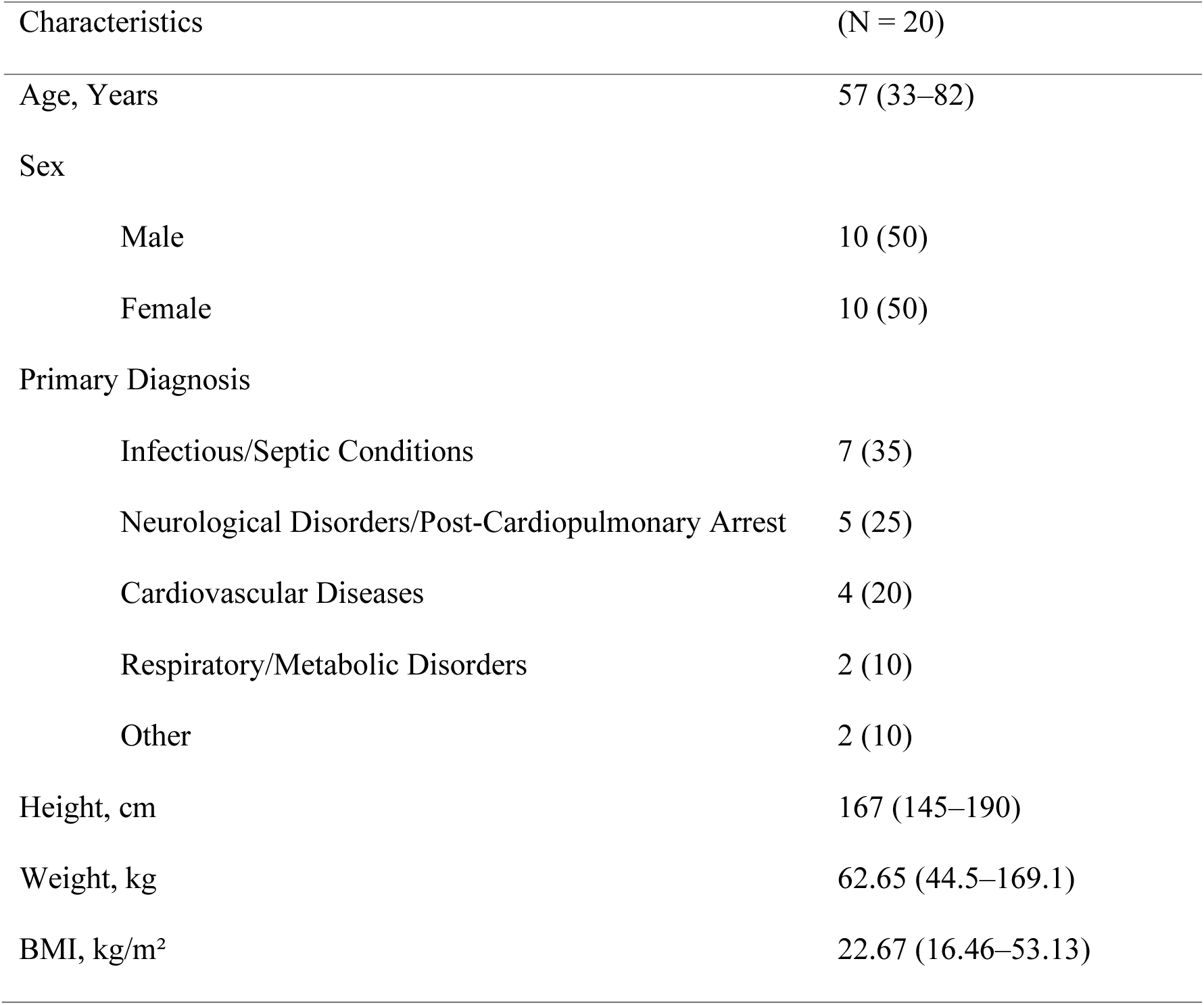
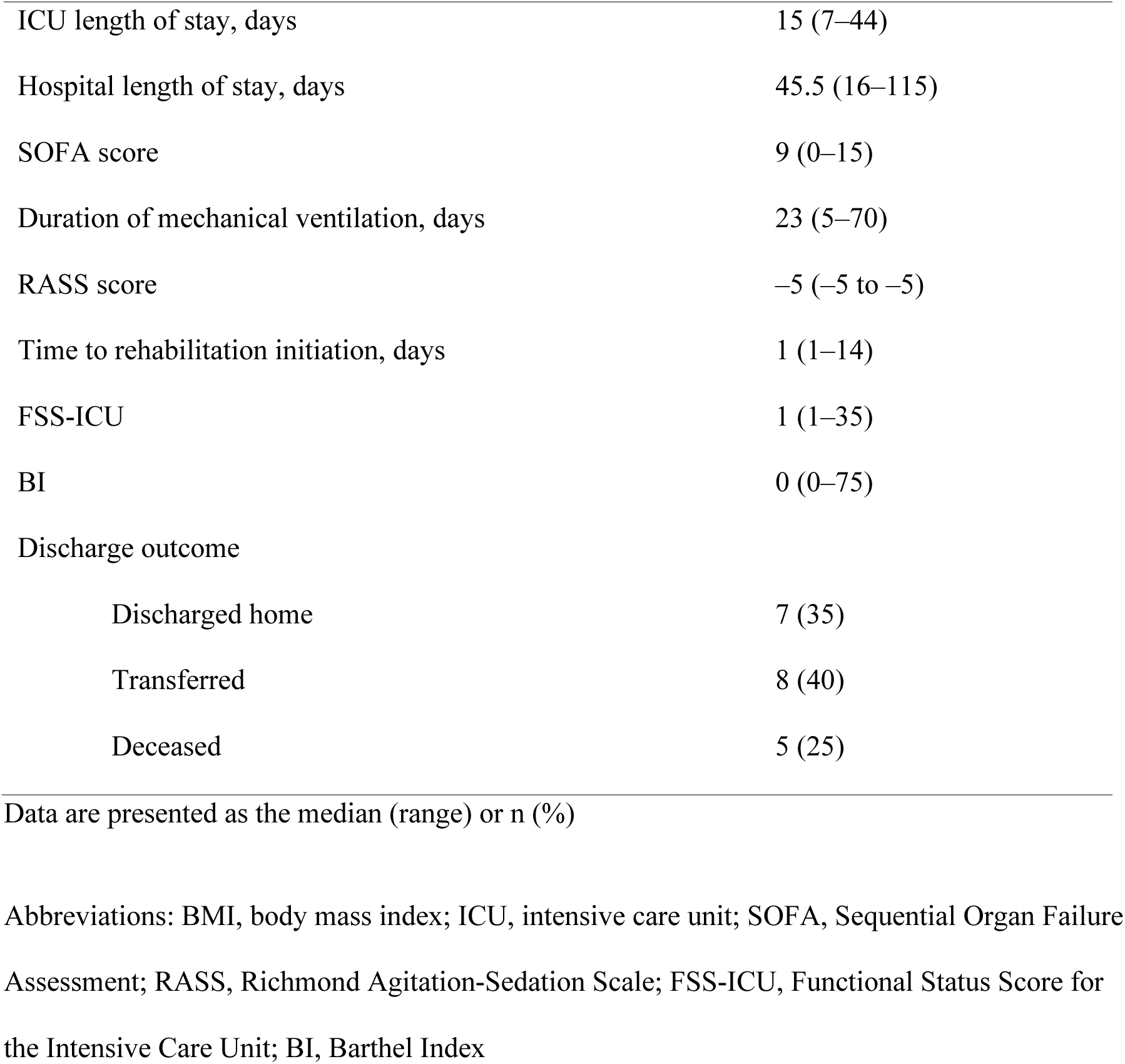
Participant Characteristics.

| Characteristics | (N = 20) |
| --- | --- |
| Age, Years | 57 (33–82) |
| Sex |  |
| Male | 10 (50) |
| Female | 10 (50) |
| Primary Diagnosis |  |
| Infectious/Septic Conditions | 7 (35) |
| Neurological Disorders/Post-Cardiopulmonary Arrest | 5 (25) |
| Cardiovascular Diseases | 4 (20) |
| Respiratory/Metabolic Disorders | 2 (10) |
| Other | 2 (10) |
| Height, cm | 167 (145–190) |
| Weight, kg | 62.65 (44.5–169.1) |
| BMI, kg/m<sup>2</sup> | 22.67 (16.46–53.13) |
| ICU length of stay, days | 15 (7–44) |
| Hospital length of stay, days | 45.5 (16–115) |
| SOFA score | 9 (0–15) |
| Duration of mechanical ventilation, days | 23 (5–70) |
| RASS score | –5 (–5 to –5) |
| Time to rehabilitation initiation, days | 1 (1–14) |
| FSS-ICU | 1 (1–35) |
| BI | 0 (0–75) |
| Discharge outcome |  |
| Discharged home | 7 (35) |
| Transferred | 8 (40) |
| Deceased | 5 (25) |

Data are presented as the median (range) or n (%)
Abbreviations: BMI, body mass index; ICU, intensive care unit; SOFA, Sequential Organ Failure Assessment; RASS, Richmond Agitation-Sedation Scale; FSS-ICU, Functional Status Score for the Intensive Care Unit; BI, Barthel Index

### Temporal Changes in Muscle CSA

The means and standard deviations of the FCU and BB CSAs at each time point are shown in Table 4. On day 0, the CSA of the FCU was 1.36 ± 0.46 cm², decreasing to 1.19 ± 0.42 cm² on day 5, 1.11 ± 0.39 cm² on day 7, and 1.08 ± 0.40 cm² on day 14. A similar trend was observed for the BB CSA values (day 0, 6.08 ± 2.54 cm²; day 5, 5.69 ± 2.73 cm²; day 7, 5.71 ± 2.60 cm²; and day 14, 5.09 ± 2.40 cm²). As shown in Table 4, the relative changes in CSA of the FCU and BB, calculated based on the values obtained on day 0, indicated a decrease in CSA in both muscles during the first 14 days after ICU admission.

**Table 4.** Temporal Changes in Muscle Cross-Sectional Area.

| Muscle | Day 0 | Day 5 | Day 7 | Day 14 | Day 5 | Day 7 | Day 14 |
| --- | --- | --- | --- | --- | --- | --- | --- |
| | (Mean ±<br>SD) | (Mean ±<br>SD) | (Mean ±<br>SD) | (Mean ±<br>SD) | (Mean<br>$\Delta\%$) | (Mean $\Delta\%$) | (Mean $\Delta\%$) |
| FCU | $1.36 \pm 0.46$ | $1.19 \pm 0.42$ | $1.11 \pm 0.39$ | $1.08 \pm 0.40$ | -12.50% | -18.38% | -20.59% |
| BB | $6.08 \pm 2.54$ | $5.69 \pm 2.73$ | $5.71 \pm 2.60$ | $5.09 \pm 2.40$ | -6.41% | -6.09% | -16.28% |

Abbreviations: SD, standard deviation; FCU, flexor carpi ulnaris; BB, biceps brachii

Friedman tests showed significant changes in CSA over time for both the FCU (χ²(3) = 22.38, p < 0.01) and BB (χ²(3) = 22.38, p < 0.01). Based on these findings, Wilcoxon signed-rank tests were performed for pairwise comparisons. For the FCU, significant reductions in CSA were observed between day 0 and day 5 (p < 0.001), day 0 and day 7 (p < 0.001), day 0 and day 14 (p < 0.001), and day 5 and day 14 (p < 0.001) (Figure 2a). For the BB, significant reductions were observed between day 0 and day 14 (p < 0.001), day 5 and day 14 (p < 0.001), and day 7 and day 14 (p < 0.001) (Figure 2b).

**Figure 2.**
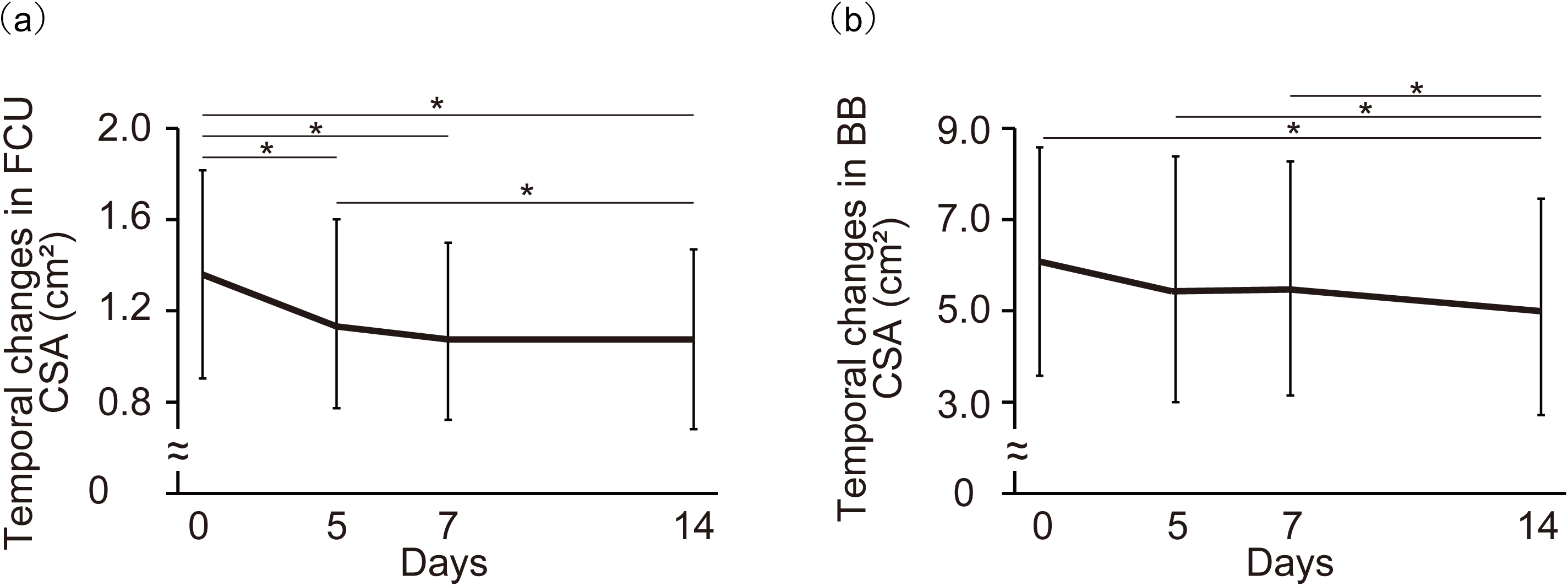
Temporal Changes in the FCU and BB CSAs. (a) The FCU CSA was measured at days 0, 5, 7, and 14. (b) The BB CSA was measured at days 0, 5, 7, and 14. Abbreviations: CSA, cross-sectional area; FCU, flexor carpi ulnaris; BB, biceps brachii; ICU, intensive care unit *(p < 0.05)

### Correlation Between the CSAs of the FCU and BB

A significant positive correlation was observed between the CSA of the FCU and that of the BB at day 0 (Spearman’s ρ = 0.8447, p < 0.001) (Figure 3a). Additionally, a moderate positive correlation was observed between the relative percentage changes in CSA from day 0 to day 14 (Spearman’s ρ = 0.5489, p = 0.0122) (Figure 3b).

**Figure 3.**
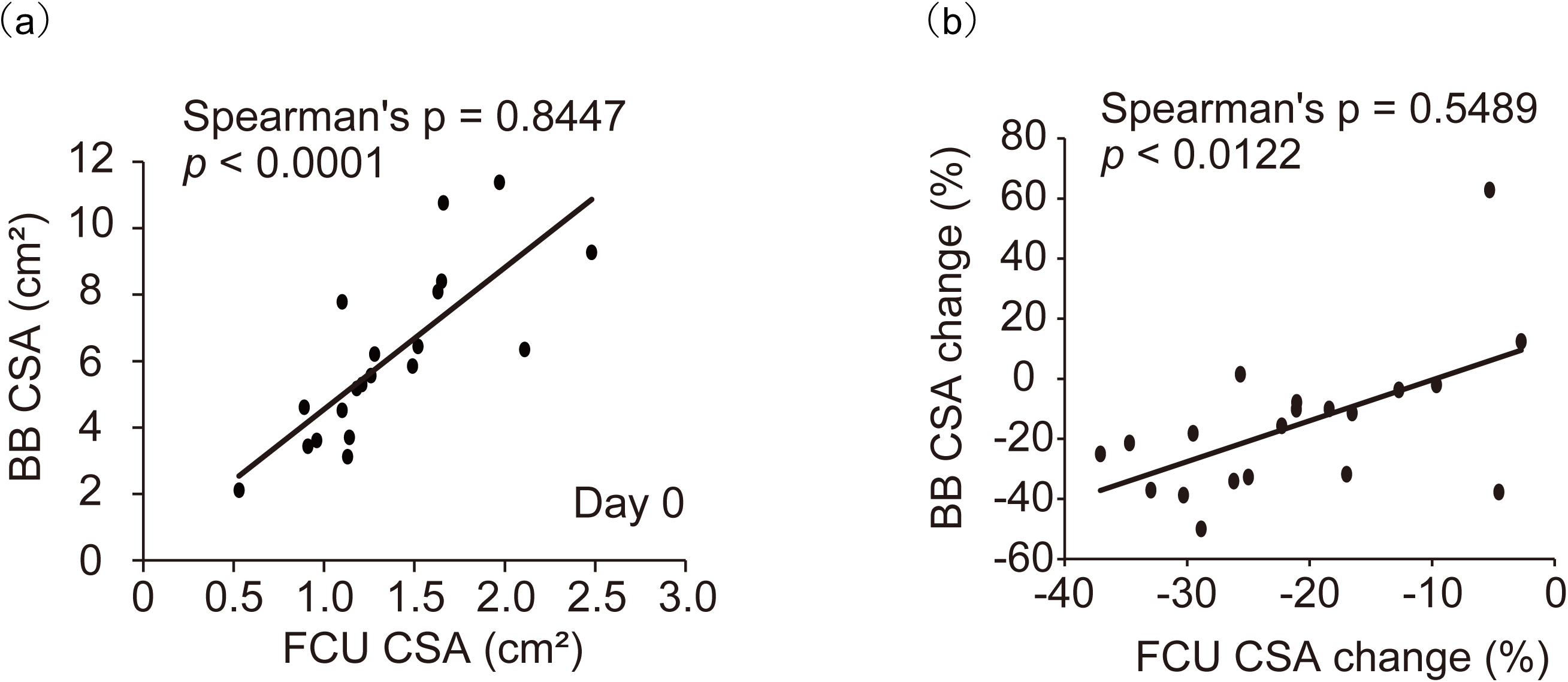
Correlation Between the CSAs of the FCU and BB. (a) Scatter plot showing the relationship between the FCU CSA and BB CSA at day 0. (b) Scatter plot showing the correlation between the changes in the FCU CSA (%) and BB CSA (%) over 14 days. Abbreviations: CSA, cross-sectional area; FCU, flexor carpi ulnaris; BB, biceps brachii

### SEM of Factors Affecting the FCU and BB CSA Changes

SEM showed that the percentage change in the BB CSA (BB 0–14-day change%) had a significant positive effect on the percentage change in the FCU CSA (FCU 0–14-day change%) (standardized β = 0.249, p = 0.0011). Moreover, BMI was observed to significantly influence the initial FCU CSA (FCU0) (standardized β = 0.042, p = 0.0004). In contrast, clinical background factors such as age, SOFA score, and duration of mechanical ventilation showed no significant effect on the FCU 0–14-day change% (all p > 0.05). Furthermore, FCU0 itself was not a significant predictor of the FCU 0–14-day change% value (p = 0.7115) (Figure 4, Table 5).

**Figure 4.**
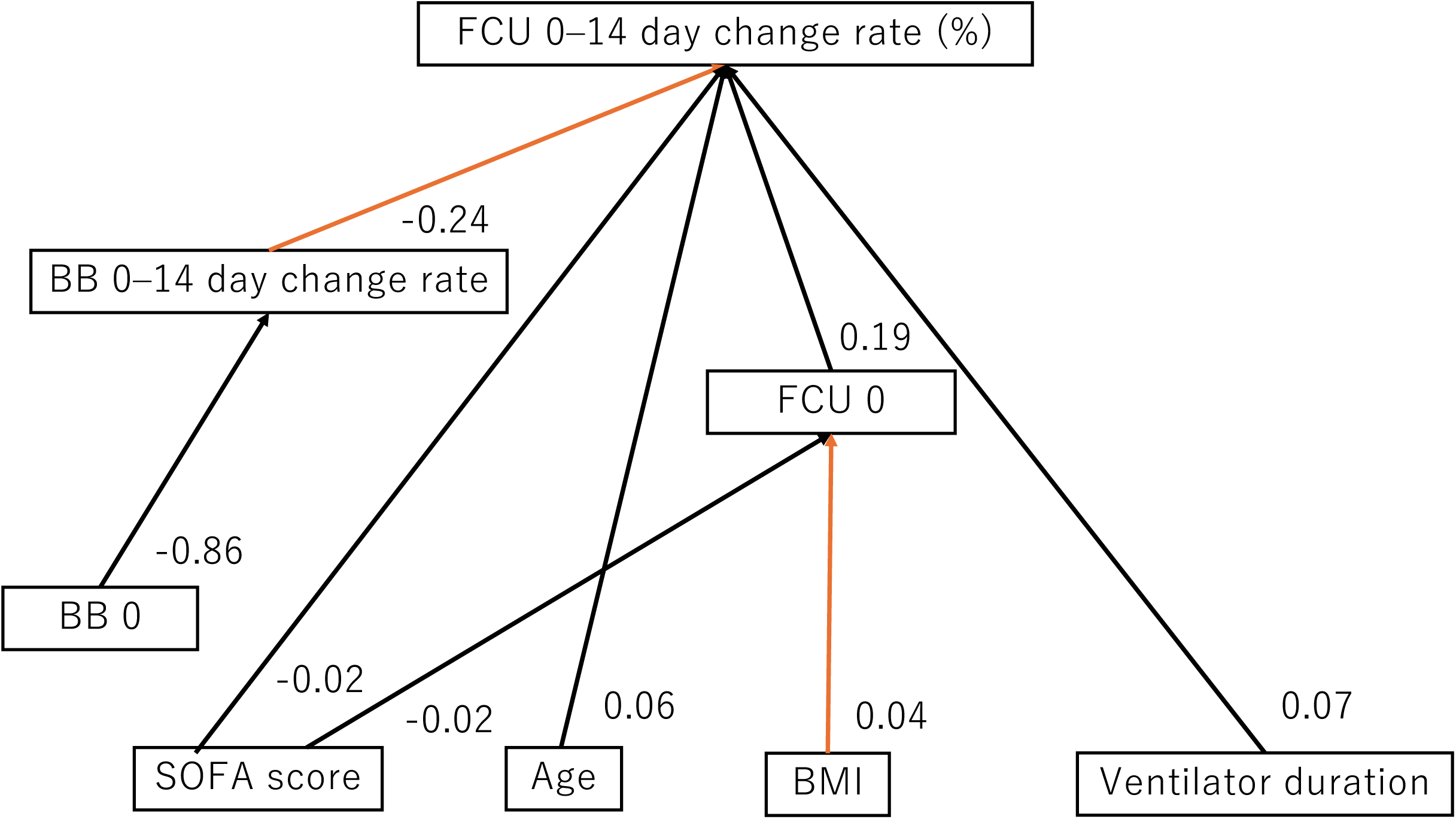
Path Diagram Showing Predictors of the FCU CSA Change Rate in ICU Patients. Solid arrows indicate statistically significant paths (p < 0.05), while dashed arrows indicate non-significant paths. Significant positive effects were observed between the following factors: from the BB CSA change rate to the FCU CSA change rate (β = 0.249) and from BMI to the FCU0 (β = 0.042). Other clinical variables—including age, SOFA score, ventilator duration, and the FCU0—did not significantly predict the FCU CSA change. Abbreviations: FCU, flexor carpi ulnaris; CSA, cross-sectional area; BB, biceps brachii; SOFA, Sequential Organ Failure Assessment; BMI, body mass index; FCU0, baseline flexor carpi ulnaris cross-sectional area (on day 0); BB0, baseline biceps brachii cross-sectional area (on day 0)

**Table 5.** Structural Equation Modeling Assessment of the Predictors of the FCU CSA Change Rate.

| Path (Predictor → Outcome) | Estimate ($\beta$) | p-value |
| --- | --- | --- |
| BB 0–14-day change rate (%) → FCU 0–14-day change rate (%) | 0.2488 | 0.0011 |
| BMI → FCU0 | 0.0416 | 0.0004 |
| SOFA score → FCU0 | −0.0295 | 0.1942 |
| BB0 → BB 0–14-day change rate (%) | −0.8614 | 0.6882 |
| Age → FCU 0–14-day change rate (%) | 0.0666 | 0.6334 |
| SOFA score → FCU 0–14-day change rate (%) | −0.0269 | 0.6223 |
| Ventilator duration → FCU 0–14-day change rate (%) | 0.0742 | 0.5085 |
| FCU0 → FCU 0–14-day change rate (%) | 0.1918 | 0.7115 |

Abbreviations: BB, biceps brachii; FCU, flexor carpi ulnaris; BMI, body mass index; SOFA, Sequential Organ Failure Assessment; FCU0, baseline flexor carpi ulnaris cross-sectional area (on day 0); BB0, baseline biceps brachii cross-sectional area (on day 0)

## Discussion

In this study, we investigated the potential of the FCU CSA as a novel and practical index for early assessment of muscle mass in critically ill patients admitted to the ICU—in comparison with the BB, which has traditionally been used in CSA evaluations. Specifically, we longitudinally measured the FCU CSA using ultrasound and examined its correlation and causal relationship with the BB CSA using SEM.

Our results revealed a significant positive correlation between the FCU and BB CSAs, and SEM analysis demonstrated that changes in the BB CSA significantly influenced changes in the FCU CSA. These findings suggest that the FCU CSA reflects the BB muscle mass and atrophic tendencies to a certain degree, indicating its potential utility as an alternative index for muscle mass assessment in the ICU setting. Moreover, Friedman test analyses showed significant reductions in CSA over time for both the FCU and BB from day 0 to day 14. Notably, the FCU exhibited a 12.5% decrease as early as day 5 and a 20.6% decrease by day 14, while the BB showed a 16.3% reduction by day 14. These findings suggest that the FCU may sensitively detect early muscle atrophy progression, potentially aiding decisions regarding earlier intervention.

Our BB CSA findings align with those of previous studies, such as those of Nakanishi et al. [8], who reported a 16.9% decrease in the BB CSA within 7 days after admission to the ICU. The consistency of these results with our 14-day decline of 16.3% supports the validity of our measurements. Furthermore, we found a moderate positive correlation between the relative CSA change rates of the FCU and BB from day 0 to day 14. This suggests that muscle atrophy during stays in the ICU is likely a systemic pathological process rather than localized disuse, progressing concurrently across different muscles. Despite anatomical and innervational differences between the FCU and BB, both exhibited similar atrophy patterns, which was noteworthy.

Previous studies using non-human primates have shown that the FCU contains a higher proportion of Type II (fast-twitch) muscle fibers [18], and compared to the BB—which is involved in antigravity activities—it may be more susceptible to disuse-induced early atrophy [19]. These physiological characteristics likely contribute to the FCU’s sensitivity in detecting early changes in muscle mass, supporting its utility as a reliable early indicator of ICU-acquired weakness (ICU-AW).

While muscle thickness and CSA are both commonly used in ultrasound-based muscle assessments, each has its advantages and limitations. Although muscle thickness is convenient and rapid to measure, muscle CSA provides a more comprehensive reflection of muscle volume; in fact, Franchi et al. previously reported that CSA better represents muscle mass changes than muscle thickness [20]. In our study, both the FCU and BB changes were effectively captured using CSA, reinforcing its utility as an objective early method of evaluation for ICU-AW. We also found that BMI significantly influenced the initial FCU CSA, suggesting that body composition and nutritional status play important roles in muscle mass upon admission to the ICU. As BMI is easily obtainable in clinical settings, integrating it with CSA measurements may lead to more accurate evaluations.

Notably, we found that the initial FCU CSA (FCU0) did not significantly predict subsequent CSA changes, implying that the initial muscle size alone does not determine the progression of muscle atrophy. Muscle wasting is a multifactorial condition influenced by inflammatory responses, nutritional deficits, pharmacologic interventions, organ dysfunction, and lack of mobility [6, 7, 21, 22]. Therefore, future studies should incorporate variables such as inflammatory markers, nutritional indices, and medication history to build multifactorial models for improved prediction accuracy.

While our study results provide valuable insights, several limitations should be acknowledged. First, it was conducted across two medical centers, and differences in patient populations and treatment protocols may have affected external validity. Therefore, larger multicenter studies with standardized protocols are required to improve generalizability. Second, although CSA was used as a quantitative measure, we did not directly compare it with the MRC score—commonly used to diagnose ICU-AW. Nonetheless, the correlation between the FCU and BB CSA changes suggests that the FCU may reflect atrophic trends in other muscle groups, and future studies should explore its relationship with clinical muscle strength assessments. Third, we only assessed CSA, without incorporating muscle thickness, echotexture, or electrophysiological data. Therefore, a more comprehensive evaluation combining these parameters is required to deepen our understanding of muscle atrophy.

Despite these limitations, the present study has highlighted the utility of the FCU—a forearm flexor muscle that is not conventionally evaluated—in the early quantitative assessment of muscle wasting using ultrasound-measured CSA. Given the FCU’s excellent visibility, reproducibility, and ease of measurement when compared with the upper arm muscles, it holds promise as a practical bedside screening tool.

## Conclusion

This study demonstrated that the early assessment of the FCU CSA correlates with conventional indices such as the BB CSA and reflects systemic muscle wasting trends in critically ill ICU patients. The FCU’s anatomical and physiological characteristics, including its high visibility and predominance of fast-twitch fibers, make it a suitable site for the early detection of muscle atrophy. Future research should aim to develop more comprehensive evaluation models incorporating CSA, BMI, inflammation, nutrition, pharmacologic factors, and physical activity. These findings might contribute to the establishment of practical methods for early screening and intervention planning in patients with ICU-AW.

## Data Availability

The datasets used and/or analyzed during the current study are available from the corresponding author on reasonable request.

## List of Abbreviations

BB: Biceps brachii
BI: Barthel Index
BMI: Body mass index
CSA: Cross-sectional area
FCU: Flexor carpi ulnaris
ICC: Intraclass correlation coefficients
ICU: Intensive care unit
IMS: ICU Mobility Scale
MRC: Medical Research Council
RASS: Richmond Agitation-Sedation Scale
SEM: Structural equation modeling
SOFA: Sequential Organ Failure Assessment

## Declarations

### Ethics Approval and Consent to Participate

The study was approved by the Institutional Review Board of Showa Medical University (Approval No. 2023-041-B). Verbal informed consent was obtained from each participant or their legally authorized representative (e.g., family member) once clinical stability allowed communication. The consent process was documented in the medical records.

### Consent for Publication

Not applicable.

### Authors’ Contributions

KA conceived the study, coordinated the research team, wrote the first draft of the manuscript, oversaw data integrity, and contributed to critical revision of the statistical and clinical interpretations. FK and KK conducted ultrasound assessments at both institutions and ensured protocol adherence and measurement reliability. FK and KK also assisted in data interpretation and technical manuscript review. JS and KU extracted and organized the clinical and rehabilitation data from the medical records and contributed to variable coding, data management, and drafting of the Methods section. NT, HI, and MF designed the clinical study framework, including patient eligibility criteria, ethical procedures, and data collection scheduling. NT, HI, and MF also contributed to aligning the protocol with ICU practices. AY provided supervision on statistical methodology, including sample size estimation, structural equation modeling, and figure preparation. AY also reviewed and revised the Results and Discussion sections. All authors discussed the findings, critically revised the manuscript for intellectual content, and approved the final version for submission.

## Acknowledgements

We would like to thank Editage (www.editage.jp) for English language editing. We also express our sincere appreciation to the ICU staff and rehabilitation teams at Showa Medical University Hospital and Showa Medical University Fujigaoka Hospital for their valuable support in participant recruitment and data collection.

## Authors’ Information

KA is a Ph.D. holder in Health Sciences and a licensed occupational therapist. He serves as a lecturer at Showa Medical University and specializes in neurorehabilitation and ultrasound-based muscle assessment in critically ill patients. His research focuses on bridging clinical rehabilitation with quantitative evaluation methods in the ICU.

KK, JS, and KU are occupational therapists and lecturers at Showa Medical University. They have clinical and educational expertise in rehabilitation sciences, with a focus on functional recovery and data-driven evaluation in acute and subacute settings.

FK is a physician and professor of rehabilitation medicine, with expertise in ultrasound imaging, early mobilization, and interdisciplinary rehabilitation for critically ill patients.

MF is a physician and assistant professor specializing in critical care rehabilitation and clinical protocol design, particularly in the intensive care environment.

NT and HI are physical therapists and associate professors with advanced knowledge in cardiopulmonary rehabilitation and research on early intervention strategies in ICU settings.

AY is a physical therapist and associate professor with an academic background in physiology. He specializes in biostatistics and advanced analytical methods, including structural equation modeling for rehabilitation research.

